# Early Lung Cancer Screening: A Comparative Study of CNN and Radiomics Models with Pulmonary Nodule Biologic Characterization

**DOI:** 10.1101/2024.07.06.24309995

**Authors:** Mukund Gupta, Edbert Victor Fandy, Krrish Ghindani

**Affiliations:** Prognosic.com; University of San Francisco, San Francisco, California, United States

**Keywords:** Lung Cancer Detection, Convolutional Neural Networks, Radiomics, Medical Imaging, Early Detection

## Abstract

Lung cancer has become an increasingly prevalent disease, with an estimated 125,070 deaths in the United States alone in 2024 (5). To improve patient outcomes and assist doctors in differentiating between benign and malignant pulmonary nodules, this paper developed a Convolutional Neural Network (CNN) model for early binary detection of pulmonary nodules and assessed its effectiveness compared to other approaches. The CNN model showed an accuracy of 98.47%, while the radiomics-based SVM-LASSO model and the Lung-RADS system showed accuracies of 84.6% and 72.2% respectively. This demonstrates that the CNN model is significantly more effective for the early binary detection of pulmonary nodules than both the radiomics-based model and the Lung-RADS system. The paper also discusses the applications of Deep Learning in healthcare, concluding that although AI proves to be an effective method for early lung cancer detection, more research is needed to carefully assess the role and impact of AI in healthcare.

## 1 Introduction

Lung cancer was the leading cause of cancer-related deaths globally. According to the World Health Organization (WHO), lung cancer accounts for approximately 1.8 million deaths in 2020, representing around 18% of all cancer deaths (2). The American Cancer Society estimates that in the United States alone, there will be about 234,580 new cases and 125,070 deaths from lung cancer in 2024 (5).

The cancerous characteristics that the malignant nodule exhibits can support the development of early stages of lung cancer into more advanced stages. Eventually, once it has reached later stages, such as stage III to stage IV, the cancer has already infected other parts of the body, such as nearby lymph nodes and structures in the chest, leading to metastasis. The disease’s insidious nature often results in late-stage diagnosis of malignant nodules, highlighting the crucial importance of early and regular diagnosis for improving survival rates.

One effective method for detecting lung nodules, both benign and malignant, is through CT scans. Unlike X-rays, CT scans provide highly detailed images of the lung tissue, far surpassing the resolution of standard chest X-rays. The sensitivity and high resolution of CT scans allow for the detection of very small nodules, even those less than 5 millimeters in size, that might be missed on an X-ray machine. Additionally, CT imaging can differentiate between different types of tissue, such as distinguishing between solid nodules, calcified nodules, and those containing fat or fluid. This helps in assessing the nature of the nodules and can provide clues about their benign or malignant nature without the need for invasive sampling. Low-dose CT (LDCT) scans, in particular, can be used for lung cancer screening in high-risk populations (e.g., long-term smokers). LDCT reduces radiation exposure while still providing detailed images, making both low-dose and standard CT scans effective screening tools.

This study will analyze and compare the effectiveness of a Convolutional Neural Network (CNN) model with a radiomics-based model, evaluating their effectiveness in identifying malignant pulmonary nodules. The proposed CNN model utilizes the VGG16 architecture and the AdamOptimizer to achieve high accuracy, sensitivity, and specificity in classifying lung cancer histological subtypes from CT scans. The radiomics-based model incorporates features such as intensity, shape, and texture to classify pulmonary nodules as benign or malignant. By comparing these models, we aim to determine their potential for early detection and management of lung cancer, thereby improving patient survival rates.

## 2 Literature Review

### 2.1 Biology of Pulmonary Nodules

According to the American Lung Association, a lung nodule is defined as a small mass of dense tissue on the lung (3). These nodules, also referred to as pulmonary nodules, can be either benign or malignant. Benign pulmonary nodules are non-cancerous and often result from infections, inflammatory conditions, or non-cancerous growths. Malignant nodules, on the other hand, indicate lung cancer or metastasis from other cancers.

The likelihood of a nodule being cancerous (malignant) is often related to its size, as pulmonary nodules are less than 3 centimeters in diameter. Nodules larger than this size are typically referred to as masses and may have a higher likelihood of being malignant. Other factors contributing to the formation of malignant nodules include cancer cells spreading from other parts of the body through the bloodstream or lymphatic system to the lungs, leading to metastatic cancer. However, on the contrary primary lung cancer arises when cells within the lung tissue undergo genetic mutations that cause uncontrolled growth, forming malignant nodules.

Malignant nodules tend to be larger than benign nodules, typically over 1 centimeter in diameter, although they can be smaller in the early stages. The edges of malignant nodules are usually ill-defined and blurry because the cancer cells invade surrounding lung tissue. Malignant nodules grow rapidly over time, with a shorter doubling time (the time it takes for the nodule to double in size) often between 1 and 6 months and unlike benign nodules, malignant nodules typically do not have classic benign calcification patterns (e.g., central, laminated, popcorn-like). If calcification is present, it tends to be eccentric or stippled. Malignant nodules can be located anywhere in the lung but are often found in the upper lobes. Central nodules may involve the major airways.

**Figure 1:**
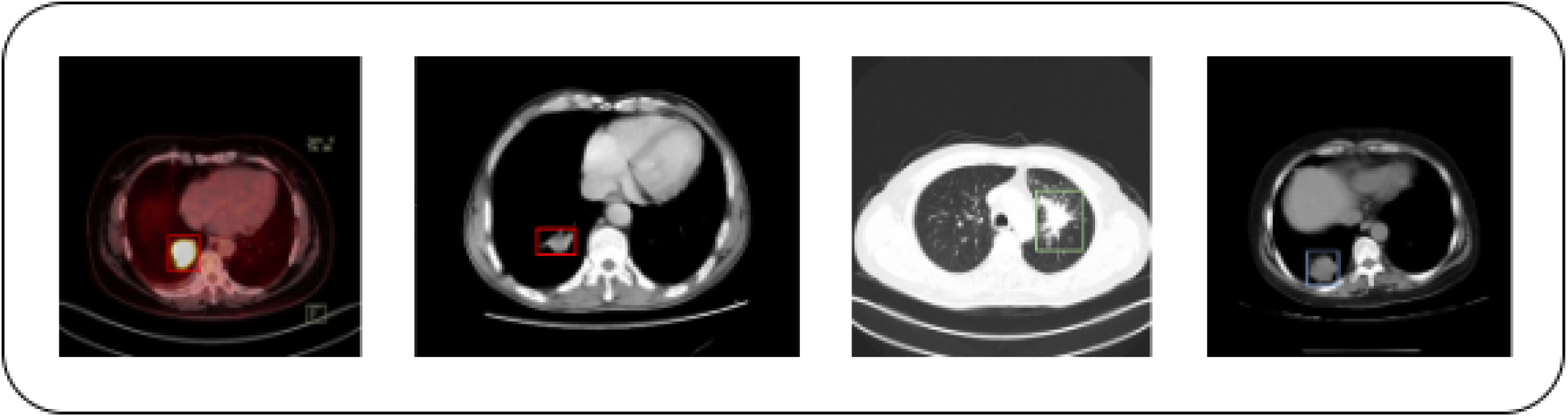
CT scan images show lung nodules with different locations and shapes.

### 2.2 CNN Model and the Current Advancements

Utilizing Convolutional Neural Network (CNN) to medical imaging research has made remarkable advancements toward early detection of lung cancer. Recent work has shown that CNNs can process complex image data for the accurate detection of malignant pulmonary nodules, as well as several subtypes of Lung Cancer: Adenocarcinoma, Squamous Cell Carcinoma, Large Cell Carcinoma, etc. Models such as VGG16 perform very well when using pre-trained feature extractors, with fine-tuning on specific tasks. CNNs have now reached sensitivity and specificity rates that are higher than those traditional feature-based methods, as well as greater order of magnitude in their ability to learn autonomously and extract relevant features from raw CT-scan images. Improvements to solutions based on continuous enhancement in the architectures of CNNs, optimization techniques, and computational power have further upgraded its performance to be an indispensable tool in medical diagnosis. Table 1 shows performance of a few classifiers on Lung CT Scan.

**Table 1:** Performance of Classifiers on Lung CT Scans.

| Cancer Type | Image Type | Classifier | Accuracy | Precision | Recall | F-Measure | Citation |
| --- | --- | --- | --- | --- | --- | --- | --- |
| Lung | CT scan | ANN | 93.3 | 91.4 |  |  | (21) |
| Lung | CT scan | MC-CNN | 87.14 | 93 |  |  | (22) |
| Lung | CT scan | CNN | 89.9 |  |  |  | (23) |
| Lung | CT scan | CNN | 92.63 | 90.7 |  |  | (24) |
| Lung | CT scan | ResNet50 + SVM (RBF) | 93.19 | 73.48 | 85.38 | 78.83 | (25) |
| Lung | CT scan | DFCNet | 89.52 | 82.54 |  |  | (26) |
| Lung | CT scan | CNN | 93.9 | 93.4 |  |  | (27) |
| Lung | CT scan | CNN | 97.9 | 98.06 | 98.07 | 98.06 | (28) |
| Lung | CT scan | EM | 96.2 | 97.4 | 98 | 98.4 | (29) |
| Proposed (Lung) | CT Scan | CNN + VGG16 | 99.56 | 99.48 | 99.61 | 99.54 |  |

**Table 2:** Performance Metrics of the Proposed CNN Model.

| Metric | Value |
| --- | --- |
| Loss | 0.0428 |
| Accuracy | 98.47% |
| Sensitivity | 95.74% |
| Specificity | 99.18% |
| AUC-ROC | 0.9981 |

## 3 Methods and Methodology

### 3.1 CNN Model Development

For the dataset, the proposed CNN model in the paper used a total of 36,300 images compiled from three separate datasets. The first and second datasets consisted of a total of 28800 infected lung images obtained from the publicly available online source “Kaggle” and the Cancer Imaging Archive (17; 15; 16; 19). Whereas, the third and last datasets consisted of 7,500 normal CT-Scans of lungs from the publicly available online source “Kaggle” and “National Cancer Institute (14; 18).

**Figure 2:**
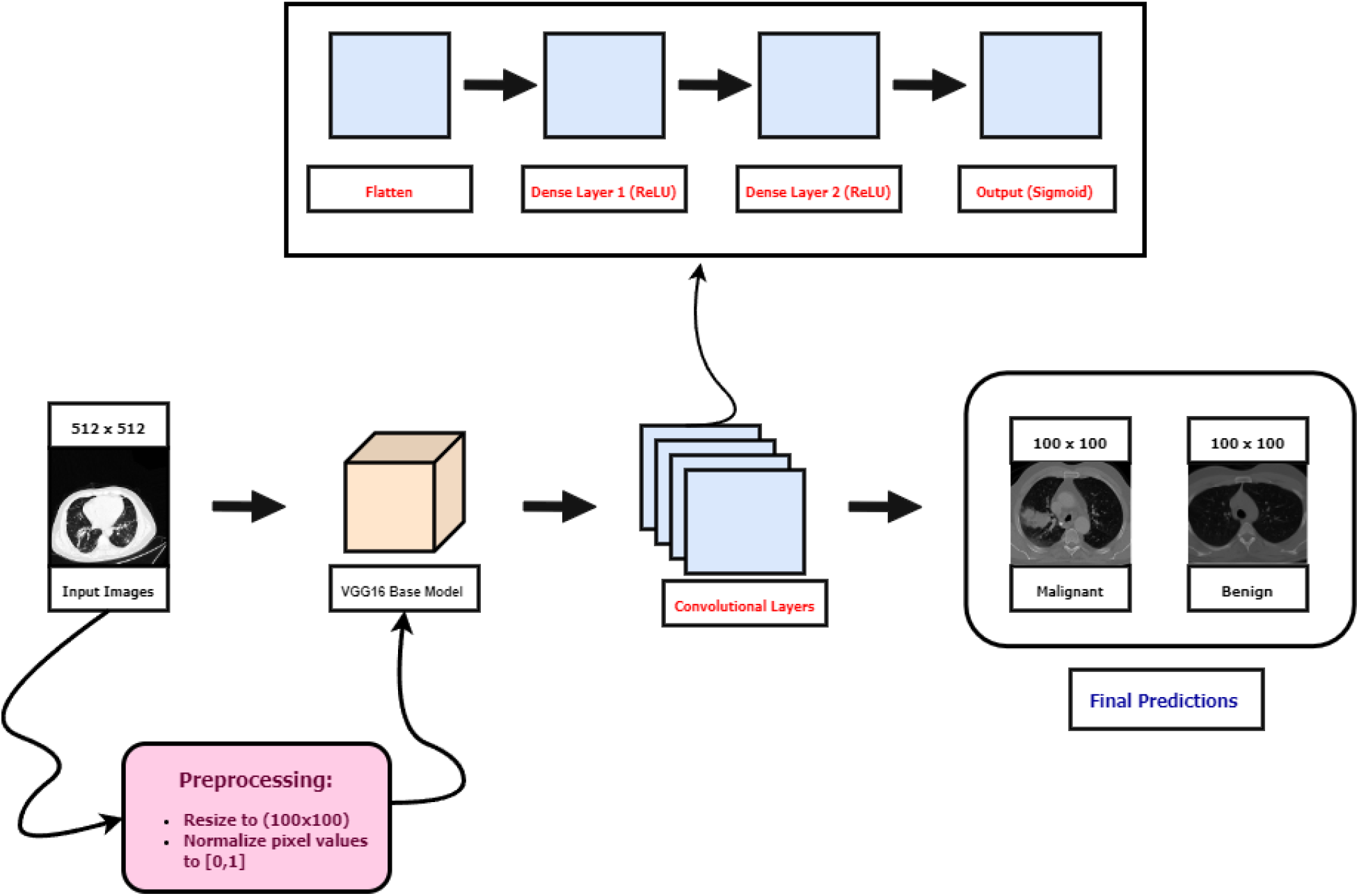
The Architecture of the CNN Model

The images were preprocessed by resizing them to (100, 100) and normalizing pixel values to the range [0, 1]. The datasets were then separated into two classes: “Infected Cases” and “Normal cases.” The dataset was split into training (80%), validation (10%), and test (10%)

The Convolutional Neural Network (CNN) model utilized the pre-trained VGG16 model as a feature extractor. The layers of the VGG16 model were then frozen to retain pre-trained weights. An additional sequential model was constructed with added dense layers for classification. The model was compiled with the Adam optimizer to minimize error functions between predicted values and actual target values. The Adam optimizer was chosen for its increased efficient convergence rates and robustness across diverse problem domains.

#### 3.1.1 Convolutional Layers

VGG16 primarily consists of convolutional layers. Given an input image matrix *I* and a convolutional filter (kernel) matrix *K*, the convolution operation is defined as:

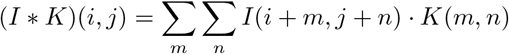

This operation involves sliding the kernel over the input image and calculating the dot product at each position. Repeating this process generates feature maps that capture local patterns in the image.

#### 3.1.2 Activation Function (ReLU)

Rectified Linear Unit (ReLU) is commonly used as the activation function after convolutional layers. The mathematical expression for ReLU is:

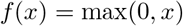

This introduces non-linearity to the model, allowing it to learn complex patterns and relationships in the data.

#### 3.1.3 Flatten Layer

The flatten layer reshapes the 3D tensor output from the convolutional layers into a 1D vector. Given an input tensor *T* of shape (*d*_1_, *d*_2_, *d*_3_), the flattening operation produces a vector *v* of length *d* _1_ *× d* _2_ *× d*_3_:

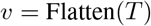

#### 3.1.4 Dense Layers

The dense (fully connected) layers perform linear transformations on the flattened feature maps. Given an input vector *x*, weight matrix *W*, and bias vector *b*, the operation is:

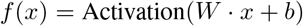

Here, the activation function (ReLU) is applied element-wise.

#### 3.1.5 Output Layer (Sigmoid Activation)

The final layer uses the sigmoid activation function for binary classification. Given a vector of raw scores *z*_*i*_, the sigmoid function is defined as:

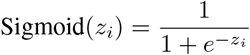

This function converts the raw scores into probability values between 0 and 1.

### 3.2 Radiomics-based Model

The model by Choi et al (1)., on the other hand, is trained to categorize the PN as benign or malignant via the extraction of quantitative features from the CT images of the nodule mechanically. The model is formulated using 103 radiomic features categorized into three groups: intensity, shape, and texture. Intensity is one of the first-order statistical measures that describe the distribution of pixel values within the PN; it contains features like mean intensity and standard deviation. Geometric shape features characterize volume, surface area, and bounding box dimensions. Texture features from the GLCM and GLRM measure spatial patterns of pixel intensities and reflect an intuition regarding tissue heterogeneity.

They used hierarchical clustering to reduce overfitting and reduce the feature space by bringing together relevant features within the unit. This is among the most powerful techniques applied to a given set of features in the entire methods discussed in this review. The correlations between similar features have been clustered, with each cluster being allowed only one representative feature. This process resulted in a refined set of features, which have much less redundancy than the initial features. In other words, they are more informatic.

A support vector machine classifier was applied with the LASSO to construct the model. SVM is a robust classification algorithm that finds an optimal hyperplane to classify different classes. LASSO adds a regularization term for the absolute size of coefficients to perform feature selection and reduce overfitting effectively. The performance of such a model was assessed using 10×10-fold cross-validation; therefore, the data set was repeatedly divided into training and validation sets. This provided a very accurate way of estimating model fit values for both in-sample and out-of-sample data.

### 3.3 Radiomics and Nodule Detection

Radiomics is the extraction of many quantitative features from medical images using sophisticated data characterization algorithms. This approach may be beneficial in analyzing subtle changes in the biology of pulmonary nodules. It possibly could be crucial for differentiating benign from malignant growths and improving early lung cancer detection.

Radiomics exploits information in LDCT scans to extract features describing the intensity, shape, and texture of the nodule. The first features are intensity features, which are the first-order statistical measures that describe pixel value distribution that exists in the nodule and include metrics like mean intensity and standard deviation.

Shape-based features define the volume of the nodule, its surface area, and the size of the minimum bounding box. Texture features are approximations of GLCM and GLRM, which are used for estimating spatial patterns of pixel intensities reflecting tissue heterogeneity.

In the model by Choi et al., 103 radiomic features are classified into intensity, shape, and texture for classifying pulmonary nodules as malignant or benign. Hierarchical clustering was used to reduce the likelihood of overfitting and increase the relevance of the features; hence, highly correlated features were grouped in the process and allowed only one representative feature from each cluster.

### 3.4 Comparison of Models

The performance of the CNN model and the radiomics-based model was evaluated using the following metrics: accuracy, sensitivity, specificity, and the area under the receiver operating characteristic curve (AUC-ROC). For the CNN model, the evaluation was conducted on a 10% test split of the LIDC-IDRI dataset (19). The cross-validation of the proposed VGG16 model was done on a separate dataset from Mendeley Data (20). This dataset comprised CT scan images collected from hospitals in Iran, including 364 images with 238 classified as cancerous and 126 as non-cancerous. The images were meticulously labeled with the assistance of a pulmonologist to ensure accurate classification and to avoid any potential errors.

In contrast, the radiomics-based model was tested using the LIDC-IDRI dataset, consisting of 72 pulmonary nodules (31 benign and 41 malignant). The radiomics-based model’s performance was further validated using additional cross-validation techniques, including 20×5-fold and 50×2-fold cross-validations, to ensure the stability and generalizability of the model.

## 4 Results

The model SVM-LASSO showed 84.6% accuracy, and the estimated AUC-ROC is 0.89. The most essential features in the prediction of malignancy generated in the classification stages are: the bounding box anterior-posterior dimension (BB_AP) and standard deviation of IDM (SD_IDM). BB_AP—this would measure how much a PN has extended in the anterior-posterior direction, and it is seen to have a very high correlation with the size of the nodule; larger nodules are more likely to be malignant. SD_IDM: texture feature—measures the directional variation in local homogeneity; since malignant nodules show more textural variation, this leads to higher SD_IDM values.

However, significant enhancement was observed in the SVM-LASSO model compared to Lung-RADS, particularly in size, type, calcification, and speculation for risk assessment. Lung-RADS achieved 72.2% accuracy with an AUC of 0.77, while the SVM-LASSO model achieved as much as 84.6% accuracy and 0.89 in AUC. The gain was statistically significant, which was indicative of the model’s superiority as a better input estimator. For that reason, this procedure was further validated with an additional 20×5-fold and 50×2-fold cross-validation to guarantee the stability of the model and generalization performance, which confirmed consistent performance with accuracies of 84.1% and 81.

**Figure 3:**
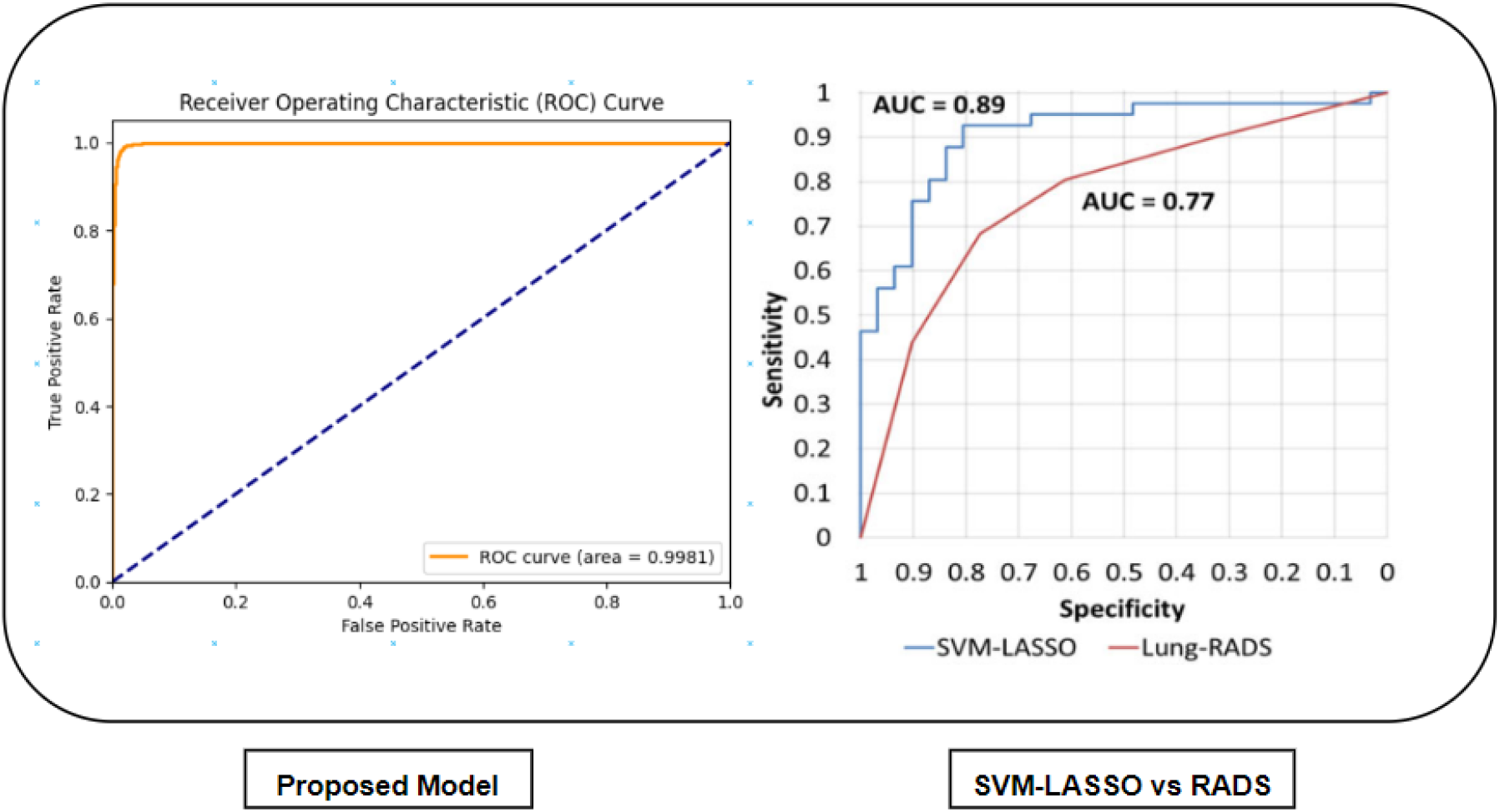
Given are the AUC-ROC Curves for the Proposed and SVM-LASSO vs Lung-RADS models respectively.

When compared to the Lung CT Screening Reporting and Data System (Lung-RADS), which primarily uses size, type, calcification, and spiculation for risk assessment, the SVM-LASSO model showed a significant improvement. Lung-RADS achieved an accuracy of 72.2% and an AUC of 0.77, whereas the SVM-LASSO model reached 84.6% accuracy and 0.89 AUC. The improvement was statistically significant, highlighting the model’s superior performance.

To ensure the model’s stability and generalizability, additional validations were performed using 20×5-fold and 50×2-fold cross-validations, which confirmed consistent performance with accuracies of 84.1% and 81.6%, respectively.

In contrast, the proposed CNN model using the VGG16 architecture showed remarkable performance metrics on the test set:

For cross-validation, the CNN model confirmed an AUC-ROC of 0.9261 on a completely unseen dataset, further demonstrating its consistent performance.

## 5 Practical Implications

Implications of AI models should continue to progress in healthcare as a means to assist medical officers in doing their tasks, for instance, a deep-learning algorithm is better at diagnosing pneumonia than radiologist, as reported in a study done by Stanford researchers (7). Regarding the ethical challenges faced in the current usages of AI in healthcare can be dealt with in accordance to the WHO’s reports published respectively in 2021 and 2024 regarding the current regulations supervising the use of AI in healthcare (8) (9).

Regulatory bodies like the FDA have also been opening the space for rigorous testing and validation of AI algorithms to detect and mitigate biases before approval (6). Although data privacy and security has become a rising concern within implications of AI in healthcare, the Health Insurance Portability and Accountability Act (HIPAA) in the U.S. and the General Data Protection Regulation (GDPR) in Europe has ensured the safety of data privacy by imposing strict regulations on data protection, assuring that AI applications comply with privacy standards (12) (13).

Other impacts that may be present as the product of the implications of AI in healthcare, such as economical impacts with rising concerns regarding the automation of tasks by AI, can lead to job displacement in the healthcare sector. A report by McKinsey Global Institute estimates that up to 800 million jobs worldwide could be displaced by automation by 2030, including healthcare roles (10). However, the report does not stop the progression of AI taking over parts of our daily lives, as the WHO emphasizes the need for workforce retraining and upskilling to adapt to AI-driven changes in healthcare. Policies such as the European Union’s Digital Education Action Plan aim to equip healthcare workers with the necessary skills for the AI era (11).

Taking all the stated things into consideration, if AI is used in healthcare worldwide, by using hosting services such as FastAPI, hospitals can easily integrate CNN models for deployment into their hospital software systems to be used by medical officers.

## 6 Conclusion

This study compared the effectiveness of a proposed Convolutional Neural Network (CNN) model and a radiomics-based model in detecting malignant lung nodules. The CNN model, using the VGG16 architecture, showed outstanding results with 98.47% accuracy, 95.74% sensitivity, 99.18% specificity, and an AUC-ROC of 0.9981. The radiomics-based SVM-LASSO model achieved 84.6% accuracy and an AUC-ROC of 0.89. The CNN model also performed well during cross-validation, with an AUC-ROC of 0.9261 on unseen data. These results highlight the CNN model’s potential to improve early lung cancer detection. However, more research is needed to refine these models, address any biases, and ensure they can be effectively used in clinical settings while maintaining ethical standards and protecting patient data.

## Data Availability

All data used in this study, including the lung CT scan images and corresponding labels, are sourced from publicly available datasets.

https://www.kaggle.com/datasets/maedemaftouni/large-covid19-ct-slice-dataset

https://www.kaggle.com/datasets/adityamahimkar/iqothnccd-lung-cancer-dataset

https://www.cancerimagingarchive.net/collection/lung-pet-ct-dx/

https://www.cancerimagingarchive.net/collection/rider-lung-ct/

https://www.cancerimagingarchive.net/collection/covid-19-ny-sbu/,

https://www.cancerimagingarchive.net/collection/lidc-idri/

